# What Can We Learn from the Time Evolution of COVID-19 Epidemic in Slovenia?

**DOI:** 10.1101/2020.05.25.20112938

**Authors:** Ioan Bâldea

## Abstract

A recent work (DOI 10.1101/2020.05.06.20093310) indicated that temporarily splitting larger populations into smaller groups can efficiently mitigate the spread of SARS-CoV-2 virus. The fact that, soon afterwards, on May 15, 2020, the two million people Slovenia was the first European country proclaiming the end of COVID-19 epidemic within national borders may be relevant from this perspective. Motivated by this evolution, in this paper we investigate the time dynamics of coronavirus cases in Slovenia with emphasis on how efficient various containment measures act to diminish the number of COVID-19 infections. Noteworthily, the present analysis does not rely on any speculative theoretical assumption; it is solely based on raw epidemiological data. Out of the results presented here, the most important one is perhaps the finding that, while imposing drastic curfews and travel restrictions reduce the infection rate κ by a factor of four with respect to the unrestricted state, they only improve the *κ*-value by ~ 15% as compared to the much bearable state of social and economical life wherein (justifiable) wearing face masks and social distancing rules are enforced/followed. Significantly for behavioral and social science, our analysis of the time dependence *κ* = *κ*(*t*) may reveal an interesting self-protection instinct of the population, which became manifest even before the official lockdown enforcement.

## 1 Introduction

In the unprecedented difficulty created by the COVID-19 pandemic outbreak,^1^ mathematical modeling developed by epidemiologists over many decades^2–7^ may make an important contribution in helping politics to adopt adequate regulations to efficiently fight against the spread of SARS-CoV-2 virus while mitigating negative economical and social consequences. The latter aspect is of paramount importance^8^ also because, if not adequately considered by governments currently challenged to deciding possibly under dramatic circumstances and formidable tight schedule, it can jeopardize the healthcare system itself. As an effort in this direction, we drew recently attention^9^ to the general fact that the spread of the SARS-CoV-2 virus in smaller groups can be substantially slowed down as compared to the case of larger populations. In this vein, the time evolution of COVID-19 disease in the two million people Slovenia certainly deserves special consideration, as on 15 May 2020, concluding that this country has the best epidemic situation in Europe, Prime Minister Janez Janska declared the end of the COVID-19 epidemic within Slovenian borders.^10^ Subsequent developments (only four new cases between 15 and 24 May,^11^ cf. Table 1) have fortunately given further support to this declaration. Attempting to understanding and learning from this sui generis circumstance is the very aim of the present paper.

**Table 1:** Total Confirmed Cases and New Confirmed Cases per Day of COVID-19 Epidemic in Slovenia Reported by the National Institute of Public Health Data According to Wikipedia.^11^

| Day of 2020 | Date | Total Confirmed | New Confirmed per Day |
| --- | --- | --- | --- |
| 64 | 03-04 | 1 | 1 |
| 65 | 03-05 | 6 | 5 |
| 66 | 03-06 | 8 | 2 |
| 67 | 03-07 | 12 | 4 |
| 68 | 03-08 | 16 | 4 |
| 69 | 03-09 | 25 | 9 |
| 70 | 03-10 | 34 | 9 |
| 71 | 03-11 | 57 | 23 |
| 72 | 03-12 | 89 | 32 |
| 73 | 03-13 | 141 | 52 |
| 74 | 03-14 | 181 | 40 |
| 75 | 03-15 | 219 | 38 |
| 76 | 03-16 | 253 | 34 |
| 77 | 03-17 | 275 | 22 |
| 78 | 03-18 | 286 | 11 |
| 79 | 03-19 | 319 | 33 |
| 80 | 03-20 | 341 | 22 |
| 81 | 03-21 | 383 | 42 |
| 82 | 03-22 | 414 | 31 |
| 83 | 03-23 | 442 | 28 |
| 84 | 03-24 | 480 | 38 |
| 85 | 03-25 | 528 | 48 |
| 86 | 03-26 | 562 | 34 |
| 87 | 03-27 | 632 | 70 |
| 88 | 03-28 | 684 | 52 |
| 89 | 03-29 | 730 | 46 |
| 90 | 03-30 | 756 | 26 |
| 91 | 03-31 | 802 | 46 |
| 92 | 04-01 | 841 | 39 |
| 93 | 04-02 | 897 | 56 |
| 94 | 04-03 | 934 | 37 |
| 95 | 04-04 | 977 | 43 |
| 96 | 04-05 | 997 | 20 |
| 97 | 04-06 | 1021 | 24 |
| 98 | 04-07 | 1056 | 35 |
| 99 | 04-08 | 1092 | 36 |
| 100 | 04-09 | 1125 | 33 |
| 101 | 04-10 | 1161 | 36 |
| 102 | 04-11 | 1188 | 27 |
| 103 | 04-12 | 1205 | 17 |
| 104 | 04-13 | 1212 | 7 |
| 105 | 04-14 | 1220 | 8 |
| 106 | 04-15 | 1248 | 28 |
| 107 | 04-16 | 1269 | 21 |
| 108 | 04-17 | 1305 | 36 |
| 109 | 04-18 | 1318 | 13 |
| 110 | 04-19 | 1331 | 13 |
| 111 | 04-20 | 1336 | 5 |
| 112 | 04-21 | 1345 | 9 |
| 113 | 04-22 | 1354 | 9 |
| 114 | 04-23 | 1367 | 13 |
| 115 | 04-24 | 1374 | 7 |
| 116 | 04-25 | 1389 | 15 |
| 117 | 04-26 | 1396 | 7 |
| 118 | 04-27 | 1402 | 6 |
| 119 | 04-28 | 1408 | 6 |
| 120 | 04-29 | 1418 | 10 |
| 121 | 04-30 | 1429 | 11 |
| 122 | 05-01 | 1434 | 5 |
| 123 | 05-02 | 1439 | 5 |
| 124 | 05-03 | 1439 | 0 |
| 125 | 05-04 | 1439 | 0 |
| 126 | 05-05 | 1445 | 6 |
| 127 | 05-06 | 1448 | 3 |
| 128 | 05-07 | 1449 | 1 |
| 129 | 05-08 | 1450 | 1 |
| 130 | 05-09 | 1454 | 4 |
| 131 | 05-10 | 1457 | 3 |
| 132 | 05-11 | 1460 | 3 |
| 133 | 05-12 | 1461 | 1 |
| 134 | 05-13 | 1463 | 2 |
| 135 | 05-14 | 1464 | 1 |
| 136 | 05-15 | 1465 | 1 |
| 137 | 05-16 | 1465 | 0 |
| 138 | 05-17 | 1466 | 1 |
| 139 | 05-18 | 1466 | 0 |
| 140 | 05-19 | 1467 | 1 |
| 141 | 05-20 | 1468 | 0 |
| 142 | 05-21 | 1468 | 0 |
| 143 | 05-22 | 1468 | 0 |
| 144 | 05-23 | 1468 | 0 |
| 145 | 05-24 | 1468 | 0 |

Thanks to long standing efforts extending over many decades, a rich arsenal of theoretical methods of analyzing epidemics exists. Most of them trace back to the celebrated SIR model^2–7^ wherein the time evolution of the numbers of individuals belonging to various epidemiological classes (susceptible (S), infected (I), recovered (R), etc) classes is described by deterministic differential equations. Unfortunately, those approaches need many input parameters^12,13^ that can often be reliably estimated only after epidemics ended,^14^ which unavoidably compromises their ability of making predictions. As an aggravating circumstance, one should also add the difficulty not encountered in the vast majority of previous studies: how do the input parameters needed in model simulations change in time under so many restrictive measures (wearing face masks, social distancing, movement restrictions, isolation and quarantine policies, etc) unknown in the pre-COVID-19 era? Estimating model parameters from data fitting in a certain time interval to make predictions can easily run into a difficulty like that described in the first paragraph of Section 2.3.

As shown below, our approach obviates the aforementioned difficulty. We will adopt a logistic growth model in a form which is different from that often employed in the past^15–19^ (see Sections 2.3 and 3 for technical details). This model is considerably simpler than SIR flavors, and already turned out to be an appealing framework in dealing with current COVID-19 pandemic issues.^9,13^ Logistic functions (see equation (2) below) were utilized for studying various problems.^20–25^ Studies on population dynamics of epidemic populations^26–32^ were also frequently based on the logistic function.

Nevertheless, as anticipated, there is an important difference between the present approach (Section 2.3) and all the other approaches of which we are aware. The latter merely justify the logistic model by the fact that recorded disease numbers followed a sigmoidal curve. Shortcomings of this standpoint are delineated in the beginning of Section 2.3. The strength of the approach presented in Section 2.3 is that we do not use data fitting. Rather, we use raw epidemiological data to validate the logistic growth and straightforwardly extract the time dependent infection rate, which is the relevant model parameter for the specific case considered and makes it possible to compare how efficient different restrictive measures act to mitigate the COVID-19 pandemic, and even to get insight significant for behavioral and social science.

## 2 Results and Discussion

### 2.1 Standard Logistic Model

To briefly remind, standard logistical growth in time *t* of an infected population *n = n*(*t*) follows an ordinary differential equation

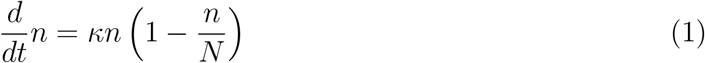

containing two constants (input model parameters): the (intrinsic) infection rate *κ*(> 0) and the so called carrying capacity *N*. In a given environment, the latter has a fixed value to which the population saturates asymptotically (lim*_t_*_→∞_ *n*(*t*) = *N*). This can be seen by straightforwardly integrating equation (1)

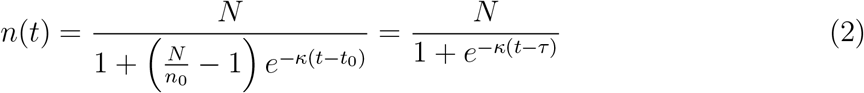

with the initial condition 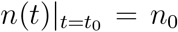, which is often recast by using the half-time 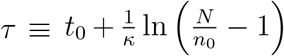, *n*(*t*)*_t=τ_* = *N*/2. Noteworthily for the discussion that follows, equation (2) assumes time-independent model parameters. In epidemiological language, *n*(*t*) gives the cumulative number of cases at time *t*. Plotted as a function of *t*, the derivative with respect to time (throughout assumed a contiguous variable) *ṅ*(*t*) ≡ *dn/dt*,

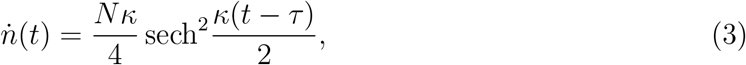

representing the “daily” number of new infections, is referred to as the epi(demiological) curve.

Figure 1a summarizes in graphical form the basic properties of logistic growth emerging the above equations. The importance of the result presented in Figure 1b will become apparent in Section 2.3.

**Figure 1:**
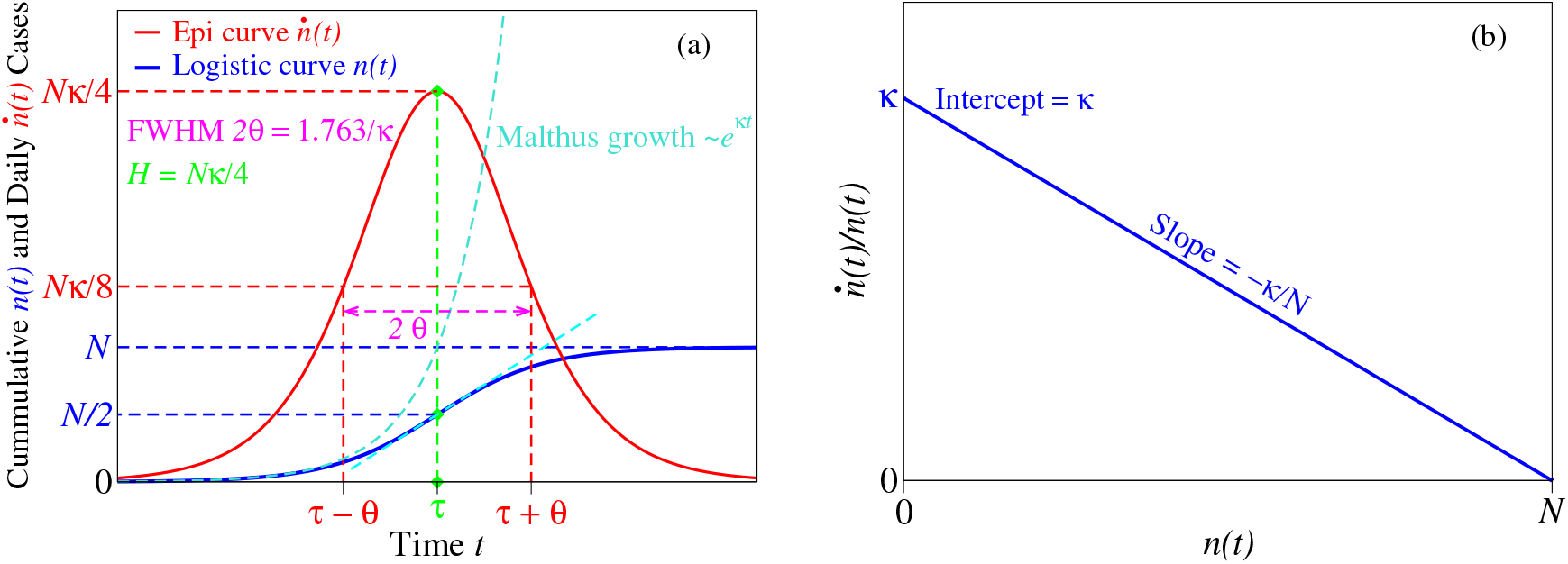
Panel a collects general properties of the standard logistic model. Panel b depicts a feature of the logistic growth whose importance is analyzed in Section 2.3.

### 2.2 Brief COVID-19 Timeline in Slovenia

Before proceeding with the data analysis let us briefly summarize relevant public heath measures, social distancing and movement restrictions imposed during the COVID-19 crisis in Slovenia.^11,33^

The first case of coronavirus was confirmed on March 4, 2020, imported via a returnee traveling from Morocco via Italy.^34^ On 10 March, the government banned all incoming flights from Italy, South Korea, Iran, and China; the land border with Italy was closed for all but freight transport; indoor public with more than 100 persons were prohibited, sporting and other events with more than 500 participants were allowed only without audience.

On 12 March, the nationwide COVID-19 epidemic was proclaimed in Slovenia. On 14 March the Crisis Management Staff of the Republic of Slovenia established by the new government led by Prime Minister Janez Jansa, confirmed on 13 March amidst the coronavirus outbreak, suspended unnecessary services. On 15 March all restaurants and bars as well as the Ljubljana Zoo were closed. On 16 March educational institutions, including kindergartens, primary and secondary schools were closed down, and public (bus, rail, air) transport was stopped. On 18 March public services (libraries, museums, cinemas, galleries were closed. On 19 March public gatherings were limited; gatherings in higher educational institutions and universities were prohibited; border checks (temperature, certificates of being healthy) were introduced. On 20 March de facto quarantine was established in Slovenia.

Significant easing occurred starting from 20 April. Border crossing gradually reopened (6, 22 and 24 April). Public transport resumed (27 April), some pupils returned to schools (27 April), all bars and restaurants as well as small hotels (up to 30 rooms) reopened for the first time since the shutdown was enforced in mid-March. On 30 April, the general prohibition of movement was lifted.

Concluding that his country has the best epidemic situation in Europe, Prime Minister Janez Janska declared on 15 May the end of the COVID-19 epidemic within Slovenian borders and allowed EU citizens free entrance.

### 2.3 Logistic Model with Time Dependent Parameters

The (blue) curve of Figure 2a depicting the evolution of total COVID-19 infections in Slovenia (underlying data are collected in Table 1) has an appealing similarity to the logistic S-shaped curve depicted in Figure 1. One would be therefore tempting to follow numerous previous authors,^26–32^ who claimed that the logistic model applies merely because of the (apparently) good data fitting.

**Figure 2:**
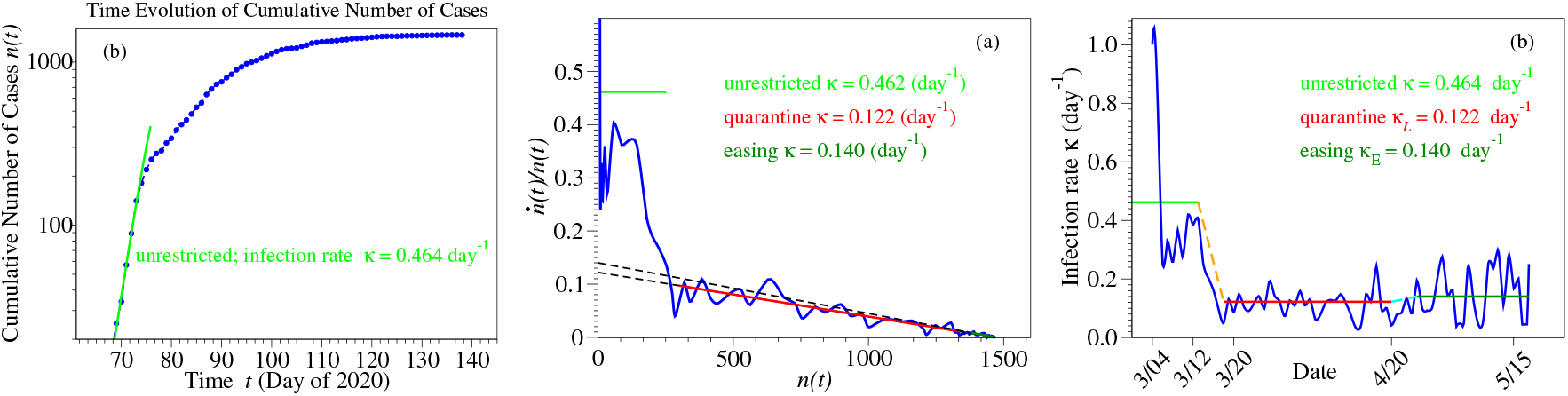
(a) In the short initial phase of the epidemic, the number of infected individuals growths exponentially in time, which allows to estimate the initial value of the infection rate (*κ* = 0.464 day^−1^. The curve of the logarithmic derivative *d* ln *n*(*t*)*/dt* = *ṅ*(*t*)*/n*(*t*) (blue line in panel b), which is merely obtained from epidemiological reports without any theoretical assumption, exhibits a linear trend that validates the description based on the logistic model. The carrying capacity *N*(= 1470) is obtained from the intersection with the x-axis. The curve of the time-dependent infection rate *κ* = *κ*(*t*) (blue line in panel c) deduced by means of equation (5) suggests a piecewise linear approximation. Noteworthy, the infection rate *κ_E_* = 0.140 day^−1^ after lockdown easing is only slightly larger than the value *κ_L_* = 0.122 day^−1^ estimated for strict lockdown.

Still, to claim that a description based on a model like that of equation (2) is valid, checking that the model parameters do not depend on the fitting range (*t*_1_, *t*_2_) is mandatory. For the specific case considered here, this means that fitting numbers of infected individuals in time range *t*_1_ < *t* < *t*_2_ should yield, within inherent statistical errors, values of *N* and *κ* independent of *t*_1_ and *t*_2_. And, like in other known cases,^35,36^ this is just the stumbling block for the logistic function approach delineated in Section 2.1. In particular, the infection rate *κ* should not depend on how broad the is the range (*t*_1_, *t*_2_); however, we checked by straightforward numerical calculations that it does.

Given the real epidemic timeline delineated in Section 2.2, the infection rate *must* indeed depend on time, *κ* = *κ*(*t*). If the contrary was true, all containment measures would be useless. But when *κ* depends on *t*, equations (2) and (3) no longer apply; they were deduced by integrating out equation (1) assuming a time-independent *κ*.

Fortunately, rather than merely inquiring how good the fitting curve based on equation (2) is, we are able to directly check (and demonstrate, see below) the validity of a *time-dependent* logistic model merely based on the real epidemiological reports. To this aim, we recast the differential equation (1), which is the basic definition of the logistic growth (*not* to be confused with the logistic function of equation (2)), as follows

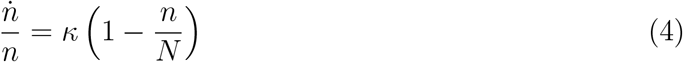

When put in this way, one can straightforwardly get insight in how to proceed. One should plot the ratio of the daily new cases to the cumulative number of cases (numerator and denominator in equation (4), respectively) as a function of the cumulative number of cases and inspect whether the curve is linear or departs from linearity. Is the decrease linear (like anticipated in the ideal simulation presented in Figure 1b), we have the demonstration that the logistic growth model applies.

The curve constructed as described above using the COVID-19 epidemic reports for Slovenia (Table 1, ref 11) is depicted in Figure 2a. As visible there, letting alone the strong fluctuations (possibly also due to the different methodology of reporting cases^11^) in the initial stage, there is a transition from a high-*κ* regime to a low-*κ* regime. Definitely, the COVID-19 restrictive measures worked. Interestingly (or, perhaps better, significantly), the low-*κ* regime appears to set in on 17-18 March (*n* = 275 − 286), suggesting a population’s prudent reaction even prior the official lockdown enforcement (20 March). This is even more important given the fact that, in view of the finite incubation time (~ 5 days), reported cases pertain to infections that occurred earlier.

An overall linear trend is clearly visible in the low-*κ* regime of Figure 2a. Equation (4) makes it then possible to estimate the carrying capacity *N* from point where the (extrapolated) straight line intersects the x-axis, while the intercept (or slope) can be used to deduce the infection rate *κ*.

Noteworthily, the low-*κ* regime comprises two periods: lockdown and lockdown easing. To quantify the differences between these two periods, we used equation (4) to analyze the epidemiological data from 18 March to 20 April (78 ≤ *t* ≤ 111) and after 20 April (*t* > 111) separately. We could not find any significant difference in *N*. With the value *N* = 1470 (to be compared with the cumulative number of cases 1468 on 24 May) in hands we have computed via equation (4) the time dependence of infection rate

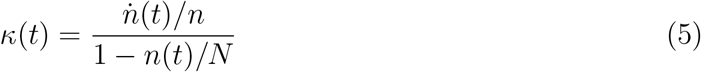

A slight difference could thus be obtained (*κ_L_* ≃ 0.122 day^−1^ versus *κ_E_* ≃ 0.140 day^−1^), leading to the schematic representation depicted in Figure 2b.

We used the piecewise linear approximation of Figure 2b for the time dependent infection rate *κ* → *κ*(*t*) to numerically integrate the differential equation (1). Results of this numerical simulations for the total and daily number of cases are depicted by the green curves in Figure 3 along with the blue curves representing the epidemiological reports.

**Figure 3:**
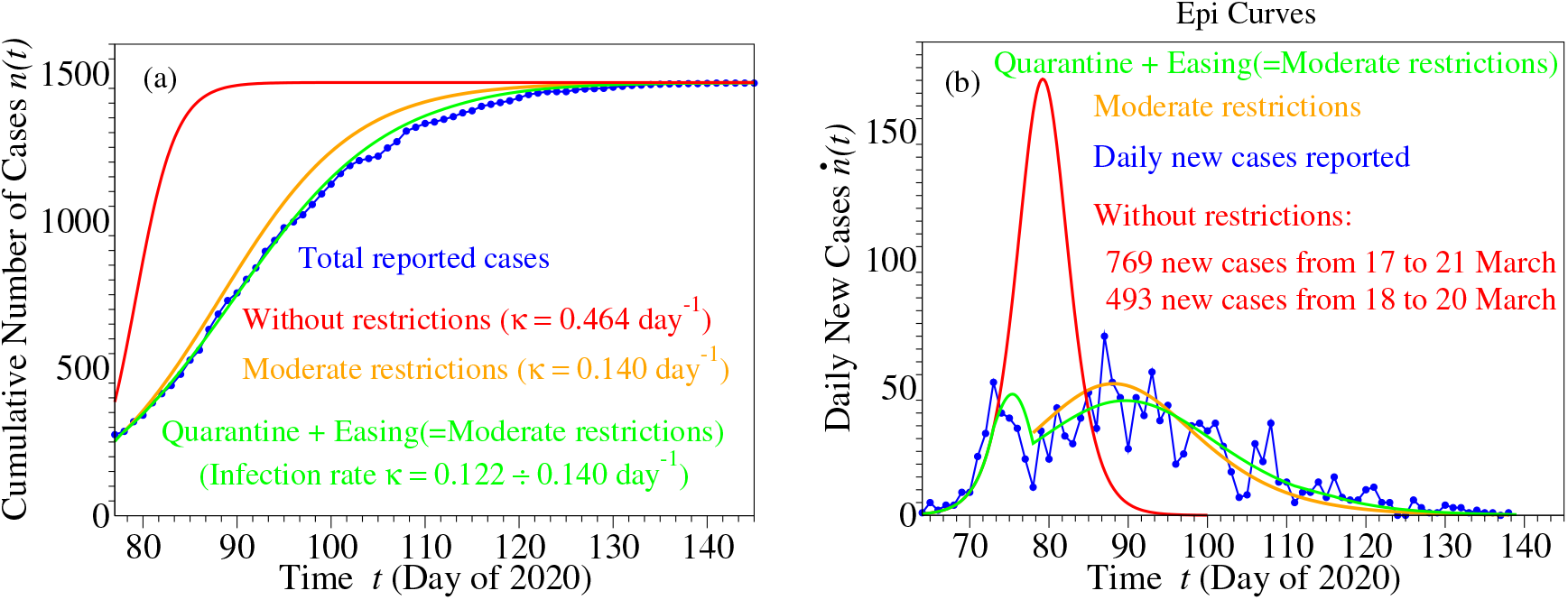
Curves for cumulative (panel a) and daily (panel b) number of cases based on epidemiological reports and simulated as described in the main text. Notice that the first peak around *t* = 73 day^−1^ does not represent a spurious effect of fluctuations. It is a real effect reproduced by numerical simulation (green curve), which nicely demonstrates how restrictive measures enforced stopped the explosive (unrestricted) evolution depicted in red. The orange curves depict results that could have been achieved if, instead of severe lockdown, the much milder restrictions effective on 20 April were imposed starting 20 March.

Time Evolution of Cumulative Number of Cases

In addition, we also show there simulations of how the Slovenian COVID-19 epidemic would have evolved if:

a. no restriction measure had been enforced. The (red) curves indicate that the result could have been grim: 769 new infections in five days or 493 new cases in three days. Again, restriction measures were definitely necessary.
b. instead of severe lockdown, much more bearable restrictive measures as effective on 20 April would have been imposed on 20 March. The rather modest differences between the orange and green curves suggest that, unless the healthcare system capacity is overwhelmed, (justified) face masks and social distancing rules can be comparably efficient in mitigating the SARS-CoV-2 virus spread with draconian lockdown while obviating paralyzing economic and social life. From this perspective, the “German model” appears to have an efficiency comparable to that of the “Italian model”.

## 3 Conclusion

We believe that especially the two results shown in Figure 2b — solely based on epidemiologically reported data without any extra (possibly speculative) theoretical supposition — are of extreme practical importance, as they could help setting adequate policies in the difficult period of the current COVID-19 pandemic:

i. Regarding the infection rate, it is worth emphasizing that, while substantially smaller than without imposing restrictions (*κ* = 0.464 day^−1^), the value *κ_L_* = 0.122 day^−1^ during shutdown is only very slightly smaller than the value *κ_E_* = 0.140 day^−1^. after easing measures effective on 20 April. The important message conveyed by this finding is that the efficiency of hardly bearable unselective quarantine and remain-isolated-at-home measures is very questionable. As one can intuitively expect, and what the present estimates do quantify, is that what really matters is not to keep everyone at home (“Italian approach”) but rather to impede virus transmission (“German approach”), e.g., by wearing masks, adequate hygiene, and social distancing. Infection transmission does not strongly increase upon easing as long as face masks and social distancing prevent SARS-CoV-2 virus spreading. One should add at this point — an important fact that appears to be currently inadequately understand — that, along with a less pleasant effect of a short-term slight increase of the daily new cases, a moderate increase in the infection rate also has a positive impact. It reduces epidemic duration; compare the *right* tail the green and orange curves in Figure 3b.
ii. The fact that the carrying capacity N does not change upon lockdown easing is equally important. This is the maximum number of individuals that can be infected in a given environment. Rephrasing, the maximum number of infected individuals does not increase when the lockdown is released; the total carrying capacity of a given environment does not change.

From a methodological perspective, one should emphasize the important technical strength of the approach proposed above, which made it possible to arrive at the aforementioned conclusions. It is only the *differential* form, equation (1), of logistic growth employed that obviates the need for any additional theoretical assumption. The traditional approach of validating the logistic model by blind data fitting using its *integral* counterpart, equation (2), does not work for COVID-19 pandemic applications because the model parameter *κ* can and does depend on time. This time dependence *κ* = *κ*(*t*) is essential to properly assess and make recommendations on the efficiency of the restriction measures to be enforced against SARS-CoV-2 virus spread.

And just because, in its differential form utilized here, the logistic model merely requires directly “measurable” epidemiological quantities (daily reports *ṅ*(*t*) and cumulative number of cases *n*(*t*), cf. equation (1)) makes in the present unsusual situation this model an alternative preferable to other more elaborate SIR-based flavors. The latter models contain a series of quantities that cannot be directly accessed “experimentally”. Governments confronted to taking decisions under unprecedented time pressure cannot await confirmation of often speculative theoretical hypotheses needed in data processing.

Before ending, let us also note that monitoring the *κ*(*t*)-timeline allowed us to get insight also relevant for behavioral and social science; the self-protection instinct of the population became manifest even before the official lockdown enforcement (cf. Section 2.3).

## Data Availability

All input data are included in the manuscript. All methods of data processing are described in the manuscript.

